# Retrospective evaluation of a filtering trabeculotomy in comparison to conventional trabeculectomy by *exact matching*

**DOI:** 10.1101/2020.01.17.20017913

**Authors:** A. Strzalkowska, P. Strzalkowski, Y. Al Yousef, J. Hillenkamp, F. Grehn, N. A. Loewen

## Abstract

**Purpose:** To compare 2-year results of a filtering trabeculotomy (FTO) to conventional trabeculectomy (TE) in open-angle glaucoma by exact matching.

**Methods:** 110 patients received an FTO and 86 a TE. FTO avoided the need for an iridectomy due to a preserved trabeculo-Descemet window anterior to the scleral flap. TE employed a trabecular block excision and iridectomy. Mitomycin C was used in both. FTO and TE were exact-matched by baseline intraocular pressure (IOP) and the number of glaucoma medications. Complete and qualified success (IOP ≤18 mmHg and IOP reduction ≥ 30%, with or without medication) were primary endpoints. IOP, visual acuity (BCVA), complications and intervention were secondary endpoints.

**Results:** 44 FTO were exact-matched to 44 TE. The IOP baseline in both groups was 22.5±4.7 mmHg on 3±0.9 medications. At 24 months, complete success was reached by 59% in FTO and 66% in TE and qualified success by 59% in FTO and 71% in TE. In FTO, IOP was reduced to 12.4±4.3 mmHg at 12 months and 13.1±4.1 mmHg at 24 months. In TE, IOP was 11.3±2.2 mmHg at 12 months and 12.0±3.5 mmHg at 24 months. Medications could be reduced at 24 months to 0.6±1.3 in FTO and 0.2±0.5 in TE. There were no significant differences between the two groups in IOP, medications, complications or interventions at any point.

**Conclusion:** Modifying aqueous flow through a limited trabeculotomy in FTO yielded clinical outcomes similar to traditional TE but allowed to avoid an iridectomy.

## Introduction

Since the first description of a guarded filtering procedure in enucleated eyes by Grant in 1958 (Grant 1958), performance of this surgery in patients in 1961 by Sugar (Saul Sugar 1961), popularization of the term “trabeculectomy” in 1968 by Cairns (Cairns 1968), and introduction of mitomycin-C as an antifibrotic to make it more effective (Chen et al. 1990), trabeculectomy (TE) has remained a primary surgery in the treatment of glaucoma (Schmier et al. 2009). Over the years, multiple modifications of this surgery have been explored to improve its effectiveness, to make outcomes more predictable and to reduce postoperative complications and need for interventions. These modifications include, among others, variations in size, localization, and thickness of the scleral-flap, different suture techniques, variable intra- and postoperative treatment with antifibrotics or a combination of these approaches with one another (Gedde et al. 2012).

In this study, we combined elements of deep sclerectomy (Hondur, Onol & Hasanreisoglu 2008, Krasnov 1968) and trabeculotomy (Esfandiari et al. 2019) with trabeculectomy in an attempt to improve conventional outflow as well as subconjunctival aqueous humor drainage. Encouraged by a pilot study of FTO with a complete success rate of 79% (Matlach et al. 2015), we hypothesized that FTO had a higher success rate and lower complication rate than TE. We applied advanced statistics, *exact matching* (Haubold & Börsch-Haubold 2017, King n.d.), to enable a highly balanced comparison of our retrospective data with 2-year follow-up.

## Materials and methods

### Study design

This retrospective study was approved by the ethics committee of the University of Würzburg, Germany. All primary open angle glaucoma (POAG) patients at the University Eye Hospital Würzburg who underwent a modified filtering trabeculotomy (FTO) with mitomycin C (MMC) or trabeculectomy (TE) with MMC by a single surgeon (FG) between 2007 and 2014 were analyzed. The indication for surgery was a failure to control IOP despite maximally tolerated medical therapy.

Patients were matched by baseline IOP and the number of glaucoma medications. We obtained the medical history, the best-corrected visual acuity assessed (BCVA [logMAR]), IOP (Goldmann tonometry [mmHg]), topical glaucoma medications (including a glaucoma medication score (GMS) (Jacobi & Krieglstein 1995)), as well as postoperative events and complications. We computed the complete and qualified success, according to the guidelines set forth by the World Glaucoma Association (Shaarawy, Sherwood & Grehn 2009). Complete success was defined as a postoperative IOP of ≤18 mmHg with a reduction of ≥30% from baseline without glaucoma medications. Qualified success was a postoperative IOP of ≤18 mmHg with a reduction of ≥30% from the baseline, achieved with or without glaucoma medications (Shaarawy, Sherwood & Grehn 2009). Follow-up visits occurred 7 days, 1 month, 3 months, 6 months, 12 months, and 24 months after surgery.

### Exclusion criteria

The exclusion criteria consisted of age below 18 years, glaucoma types other than primary open-angle glaucoma, and a history of incisional glaucoma surgery.

### Surgical technique

The eye was rotated downward with a traction suture. A 5 mm fornix-based peritomy was made at the anatomic 12 o’clock position, and a sub-tenon pocket was fashioned to accommodate a sponge soaked with mitomycin C (MMC) at a concentration of 0.2 mg/ml and 100 µl volume for 3 minutes. All patients received daily subconjunctival injections of 5 mg of 5 fluorouracil (5-FU) for one week except when IOP was at or below 5 mmHg.

In TE, a 3 mm x 4 mm half scleral thickness flap was created. A 0.8 mm x 2 mm sclerotrabecular block was excised to enter the anterior chamber, as described before (Matlach et al. 2015). A peripheral iridectomy was made. The scleral flap was secured with 10.0 nylon to allow visible percolation of aqueous, and the conjunctiva was closed with an interlocking running suture resulting in a diffusely forming bleb (Pfeiffer & Grehn 1992).

In FTO, the scleral flap was sized 4 mm x 4 mm. A smaller, tongue-shaped flap was dissected underneath to unroof and create access ostia to Schlemm’s canal. This second flap was excised similar to deep sclerectomy. The canal was probed with a metal trabeculotomy probe (Mackensen, Geuder Inc, Heidelberg) on both sides while the trabeculo-Descemet window at the base of the scleral flap was preserved so that no bulk aqueous outflow could occur and no iridectomy was needed. The remaining steps were identical to those in TE.

### Statistics

Statistical analyses were performed using Statistica 13.1 (StatSoft, Tulsa, Oklahoma, United States) and MedCalc (MedCalc 19.1.3, Ostend, Belgium). A total of 88 patients (1:1, FTO:TE) were matched with exact matching (Haubold & Börsch-Haubold 2017, King n.d.) based on the baseline IOP and glaucoma medications.

Categorical variables were described as the frequency with percentage, whereas continuous and discrete variables as mean with standard deviation (SD) or median with range. The chi-squared test or Fisher’s exact test was used for the analysis of categorical variables. Continuous variables were compared using Student’s t-test or Mann–Whitney U test, whereas discrete variables were compared using Mann-Whitney U test. The distribution of continuous variables was determined by the Shapiro-Wilk test, equality of variances by Levene’s test. Assessment of repeated measures for IOP was performed using repeated measures MANOVA and Tukey’s test, while visual acuity (logMAR) and medications were examined using the Friedman test and Wilcoxon signed-rank test. The success of treatment was expressed by a Kaplan-Meier curve and compared between treated groups using the log-rank test. The success of treatment at particular a given time point was determined by odds ratio (OR) with respective 95% confidence intervals and p-values. P-values below 0.05 were considered statistically significant.

## Results

A total of (196) patients were included. The unmatched demographic data of FTO and TE had significant differences in preoperative IOP (p=0.017), glaucoma medication score (p=0.001), and pseudophakia (p=0.012). Eighty-eight eyes (44 in each group) could be matched as exact pairs eliminating key differences in IOP and medications. The IOP at baseline was 22.6±4.7 mmHg in FTO and 22.6±4.7 in TE while on 3.0±0.9 medications in both (Table 1). There were no significant differences between FTO and TE in gender, age, best-corrected visual acuity, type of glaucoma, operated side, number of preoperative surgeries, including lens surgery or laser treatments.

**Table 1:** Demographic data of both groups after matching.

|  | **FTO n=44** | **TE n=44** | **p-value** |
| --- | --- | --- | --- |
| female (%) | 18 (41) | 21 (48) |  |
| male (%) | 26 (59) | 23 (52) | 0.520 |
| age, years (mean±SD) | 65±13 | 68±9 | 0.483 |
| BCVA, logMAR (mean±SD) | 0.2±0.3 | 0.2±0.3 | 0.923 |
| preoperative IOP, mmHg (mean±SD) | 22.5±4.7 | 22.5±4.7 | 0.891 |
| glaucoma medication (mean±SD) | 3±0.9 | 3±0.9 | 0.755 |
| type of glaucoma (%) |  |  |  |
| POAG | 33 (75) | 33 (75) |  |
| PXG | 7 (16) | 11 (25) |  |
| PG | 4 (9) | 0 (0) | 0.087 |
| right | 21 (48) | 26 (59) |  |
| left | 23 (52) | 18 (41) | 0.285 |
| pseudophakia (%) | 13 (30) | 3 (7) | 0.006 |
| prior ocular surgery including phaco (%) |  |  |  |
| 0 | 28 (64) | 40 (93) | 0.003*** |
| 1 | 15 (34) | 3 (7) |  |
| 2 | 1 (2) | 0 (0) |  |
| prior ocular surgery excluding phaco (%) |  |  |  |
| 0 | 40 (91) | 43 (100) |  |
| 1 | 4 (9) | 0 (0) | 0.116 |
| type of prior laser (%) |  |  |  |
| ALT | 13 (30) | 12 (27) | 0.813 |
| SLT | 3 (7) | 0 (0) | 0.241 |
| CPC | 4 (9) | 3 (7) | 1.000 |
| nd:YAG capsulotomy | 1 (2) | 0 (0) | 1.000 |

There were no statistically significant inter-group differences in complete or qualified success at any time (p=0.403 for complete success; p=0.204 for qualified success at 24 months, Figure 1, Supplementary Figure 1). The complete success rate in FTO ranged from 79% at 6 to 78% at 12 months and 59% at 24 months. In TE, it was 81%, 85%, and 66%, respectively. Similarly, IOPs of FTO and TE were not significantly different at any time (12 months: p=0.983, 24 months: p=1.000, Figure 2). At one year, the IOP had declined to 12.4±4.3 mmHg in FTO and 11.3±2.2 mmHg in TE with medications (qualified success). At two years, IOP remained at 13.1±4.1 mmHg in FTO and 12.0±3.5 mmHg in TE (qualified success), respectively. The postoperative visual acuity was not significantly different in FTO and TE at any time (p=0.894 after 12 months; p=0.443 after 24 months; Figure 3).

**Figure 1:**
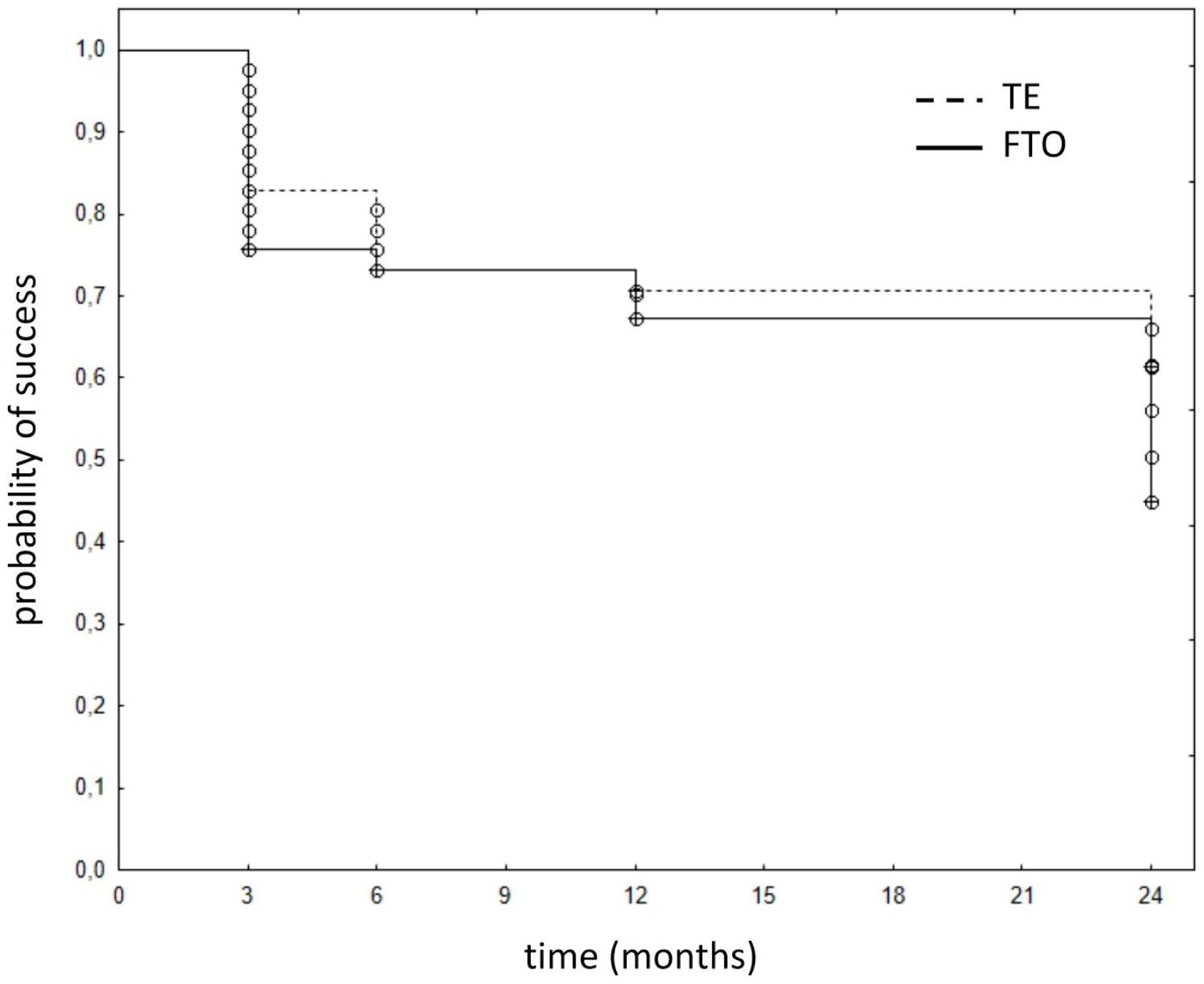
Kaplan-Meier curve for complete success in FTO and TE.

**Figure 2:**
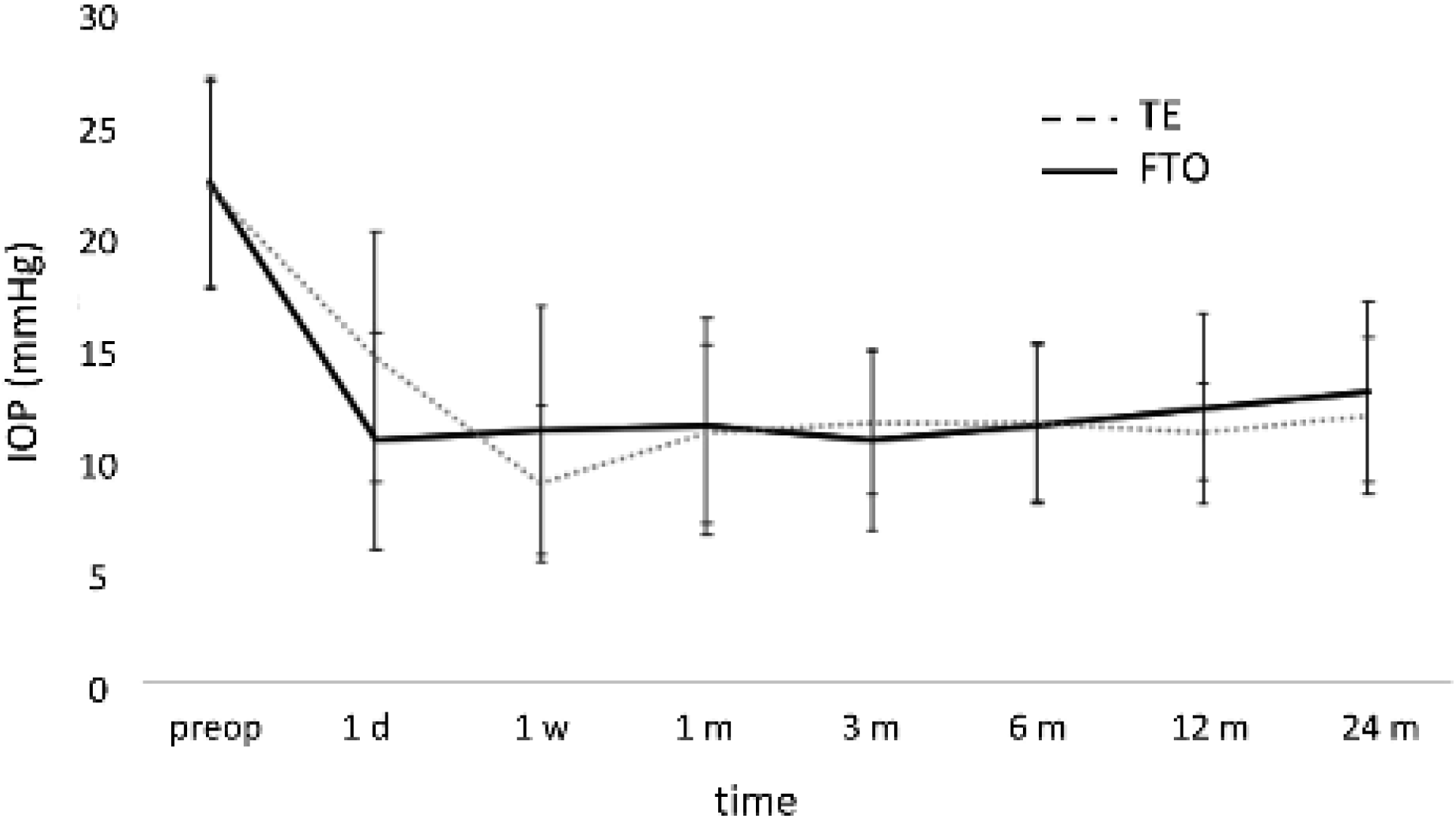
Mean preoperative vs postoperative IOP of FTO and TE.

**Figure 3:**
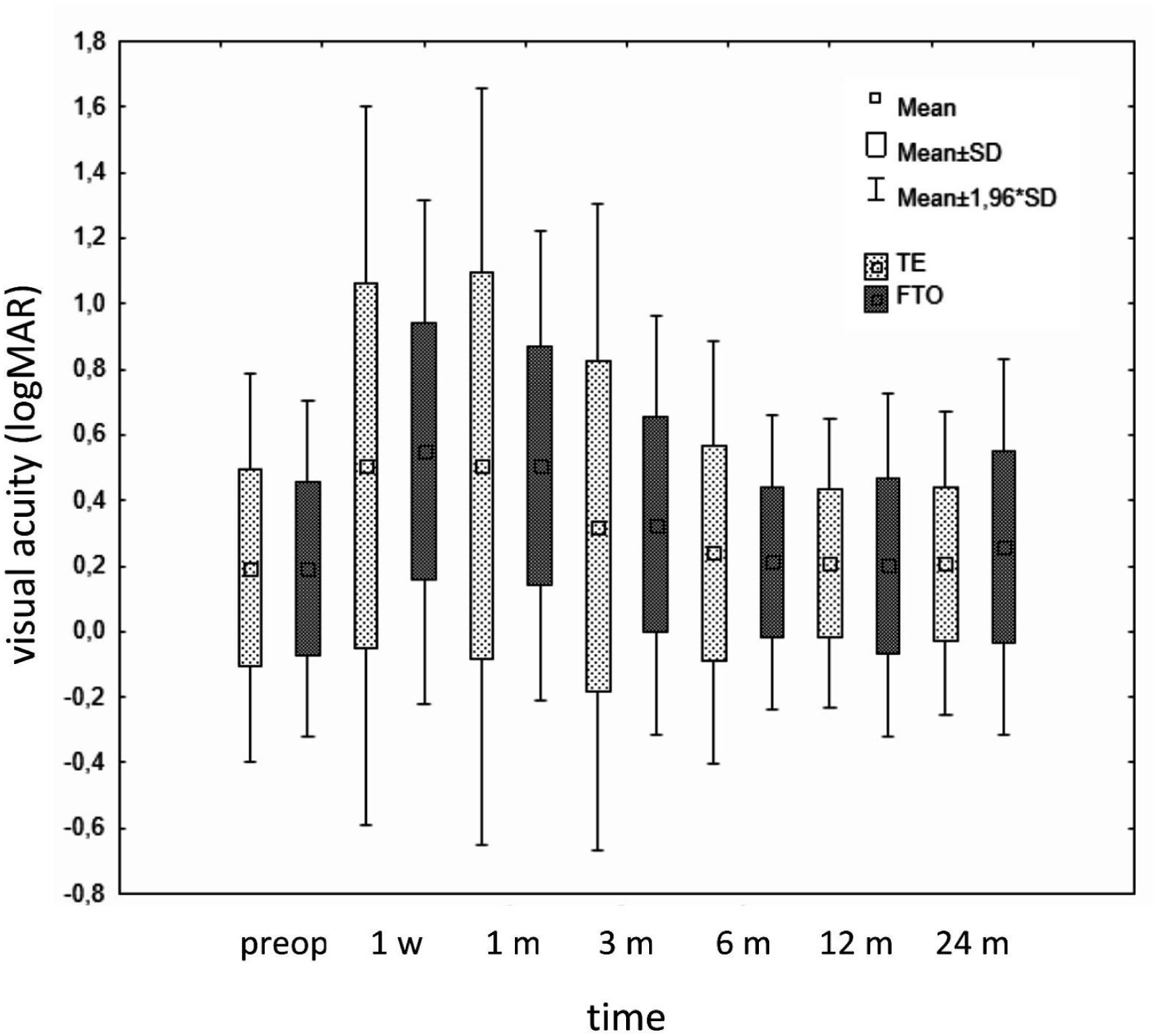
Preoperative versus postoperative visual acuity of both groups (mean±SD)

There was a reduction in glaucoma medication from 3±0.9 to 0.6±1.3 in FTO and from 3±0.9 to 0.2±0.5 in TE after 24 months. There was no significant difference between the two groups in glaucoma medications at 24 months (p=0.471) with 19% of patients in FTO and 12% of patients in TE using glaucoma drops (p=0.471, Figure 4).

**Figure 4:**
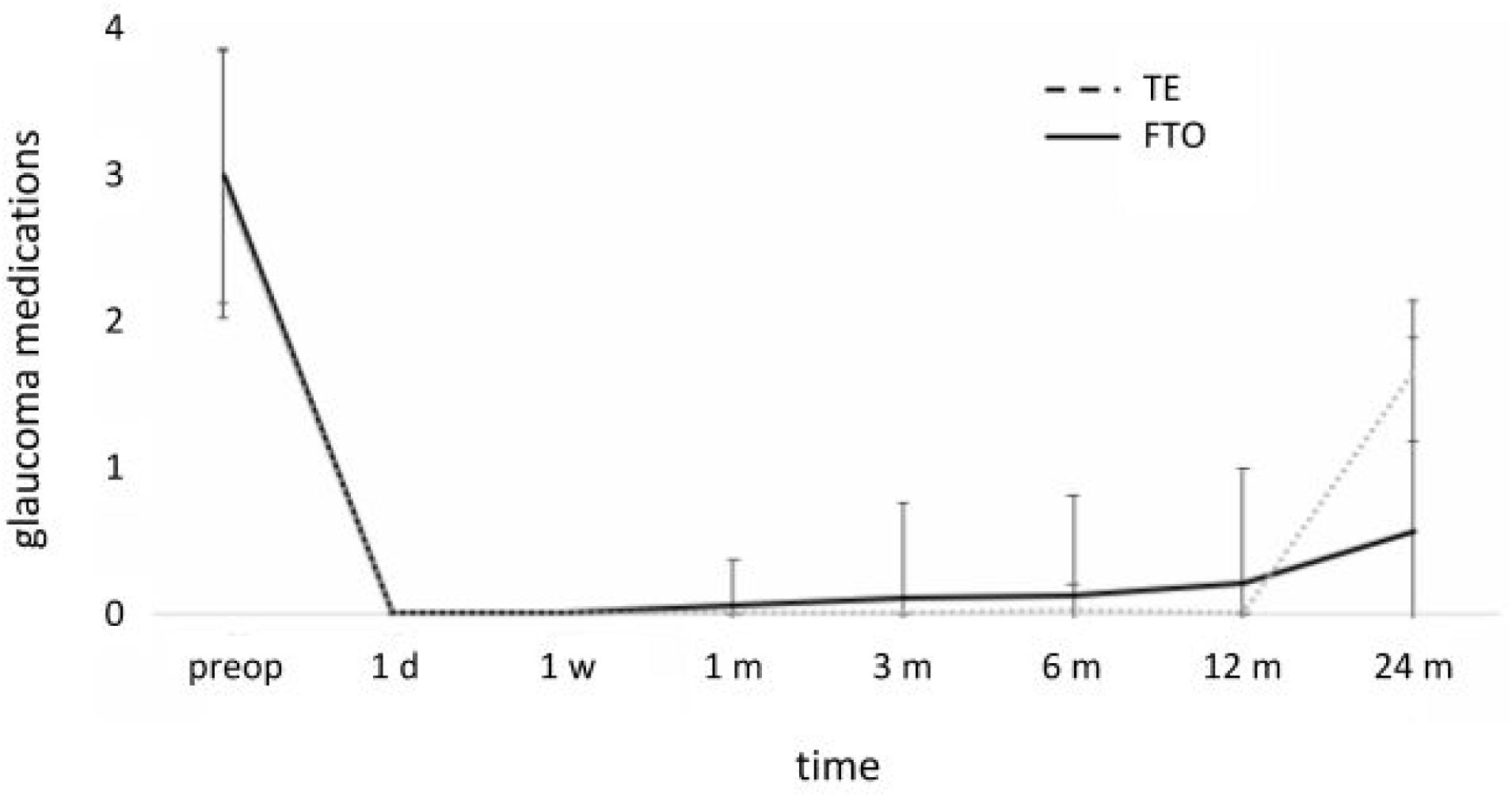
Preoperative versus postoperative glaucoma medications of both groups.

Postoperative complications in FTO included 5 eyes (12%) with a high IOP, hyphema in 5 cases (11%) and hypotony (IOP ≤ 5 mmHg (Abbas, Agrawal & King 2018)) with choroidals in 4 eyes (9%). The most common complication in TE was a flat bleb in 6 eyes (14%), hyphema in 3 (7%), high IOP in 2 eyes (5%), bleb leakage in 2 eyes (5%), and hypotony in 1 eye (2%). A total of 16 mostly reversible complications occurred in each group (37%; Table 2).

**Table 2:**
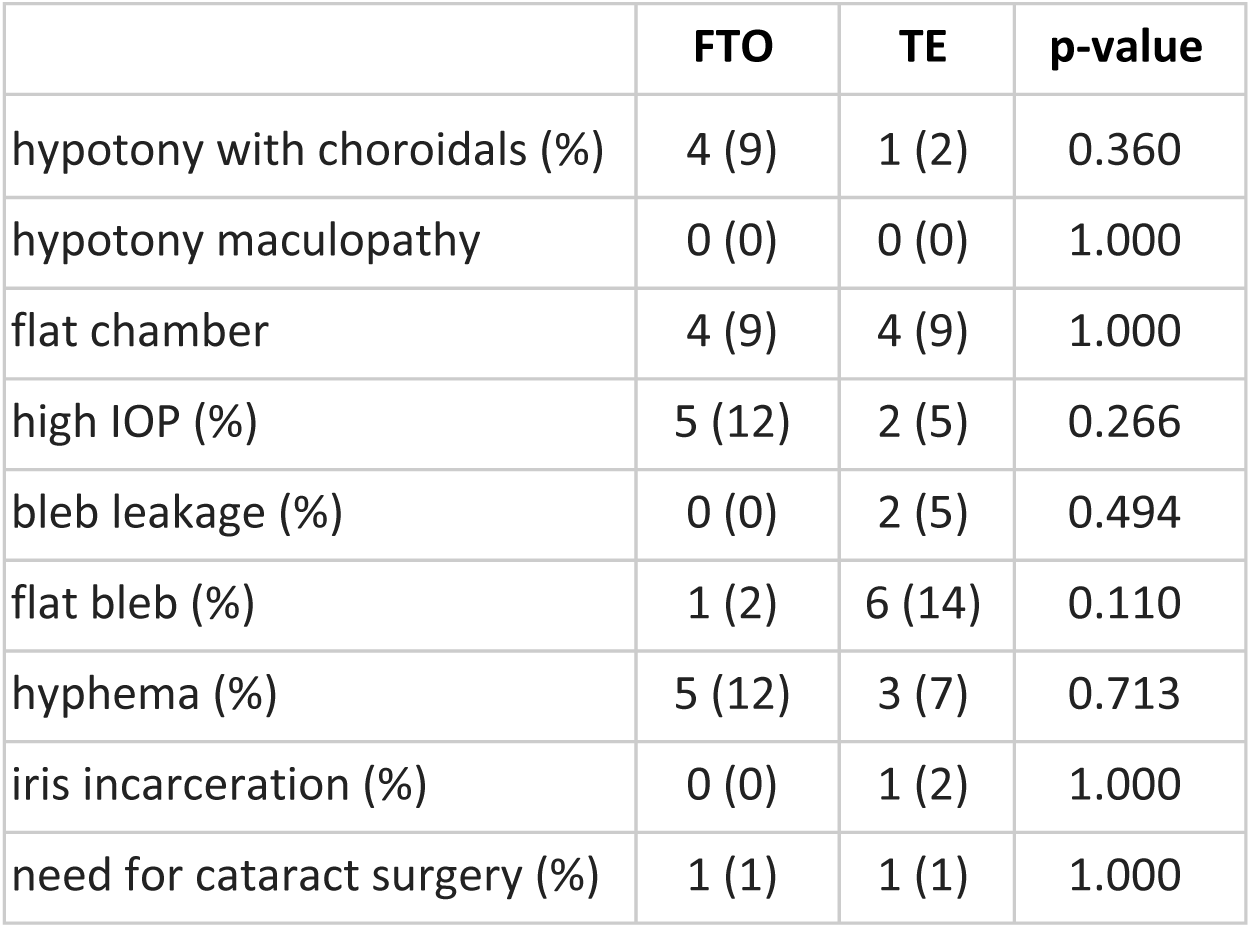
Postoperative complications of both groups.

The number of postoperative interventions was the same in the two groups (p=0.087). Bleb needling, conjunctival suture, and scleral flap suture were the most common interventions. There was no statistically significant difference between both groups in early or late interventions (Table 3), except 5-FU and laser suture lysis, which was performed more often in FTO.

**Table 3:** Postoperative interventions in FTO and TE.

| early postoperative interventions | FTO | TE | P-Value |
| --- | --- | --- | --- |
| no. of interventions |  |  |  |
| 0 (%) | 26 (59) | 35 (80) | 0.087 |
| 1 (%) | 15 (34) | 8 (18) |  |
| 2 (%) | 2 (5) | 0 (0) |  |
| 3 (%) | 1 (2) | 0 (0) |  |
| 4 (%) | 0 (0) | 1 (2) |  |
| 5-FU | 2.1±2.4 | 0.8±0.9 | 0.023 |
| bleb needling | 11 (25) | 6 (14) | 0.280 |
| conjunctival suture | 4 (9) | 3 (7) | 1.000 |
| scleral flap revision (high IOP) | 2 (5) | 0 (0) | 0.494 |
| scleral flap suture (hypotony) | 2 (5) | 2 (5) | 1.000 |
| nd:YAG laser goniotomy | 1 (2) | 0 (0) | 1.000 |
| cyclodestruction | 1 (2) | 0 (0) | 1.000 |
| iris repositioning | 1 (2) | 1 (2) | 1.000 |
| laser suture lysis (average±SD) | 1.5±1.5 | 0.8±0.9 | 0.026 |
| **late postoperative interventions** |  |  |  |
| number of interventions |  |  |  |
| 0 (%) | 35 (80) | 42 (95) | 0.147 |
| 1 (%) | 6 (14) | 2 (5) |  |
| 2 (%) | 3 (7) | 0 (0) |  |
| bleb needling | 1 (2) | 2 (5) | 1.000 |
| conjunctival suture | 3 (7) | 0 (0) | 0.241 |
| scleral flap revision (high IOP) | 3 (7) | 0 (0) | 0.241 |
| scleral flap suture (hypotony) | 0 (0) | 0 (0) | ** |
| nd:YAG laser goniotomy | 4 (9) | 0 (0) | 0.116 |
| cyclodestruction | 1 (2) | 0 (0) | 1.000 |
| Re-TE | 0 (0) | 0 (0) | ** |

\*\* not calculated

## Discussion

Trabeculectomy with MMC remains a primary surgery in the management of advanced glaucoma (Schmier et al. 2009) despite its potential for serious complications that include choroidal effusions, maculopathy, blebitis, endophthalmitis, and suprachoroidal hemorrhage (Schwartz & Budenz 2004). Numerous modifications have been explored over the years to reduce the rate of these. They include a smaller scleral flap (Ophir 2001), limbus-versus fornix-based conjunctival closure (Solus et al. 2012), releasable flap sutures (Aykan et al. 2007, Vuori & Viitanen 2001), a combination of trabeculectomy with deep sclerectomy (Kayikcioglu, Emre & Kaya 2010), different concentrations and exposure times of MMC (Bindlish et al. 2002) or sutureless tunnel trabeculectomy without iridectomy (Eslami et al. 2012). The iridectomy that is part of traditional trabeculectomy with a trabecular block excision can cause hyphema, inflammation, posterior synechiae, iridodialysis, and cataracts (de Barros et al. 2009, Grehn & Müller 1990). FTO addresses some of these issues by creating a more spread-out intake of aqueous humor, thereby reducing iris aspiration and avoiding the need for an iridectomy. The trabeculotomy and sclerectomy (Grehn 1995, Krasnov 1964, Krasnov 1968), that are part of the FTO, were meant to remove some of the post-trabecular outflow resistance (Dvorak-Theobald & Kirk 1956, McDonnell et al. 2018, Waxman et al. 2018).

We used matching, a nonparametric method of controlling the confounding influence of pretreatment variables in observational data (Blackwell et al. 2009). Before matching, FTO and TE had significant differences. We have previously used coarsened exact matching, propensity score matching, and, more recently, exact matching. Coarsened exact matching applies multiple imputations to fill in missing data as not to distort any relationships contained in the data while enabling the inclusion of all observed data from moderately uneven groups. Such was the case when we compared patients with a primary IOP indication to patients with a mixed indication for both cataract removal and IOP reduction (Honaker et al. 2011, Neiweem et al. 2016, Parikh et al. 2016). Propensity score matching is helpful to compare even more divergent groups, for instance, patients undergoing tube shunt surgery with patients undergoing trabectome surgery (Esfandiari et al. 2018, Kostanyan et al. 2017). By contrast, exact matching is well suited to compare similar pathological conditions and similar treatments, for instance, phaco-iStent or phaco-trabectome (Al Yousef et al. 2019, Esfandiari et al. 2018). A downside of exact matching is that a certain number of datasets must be excluded from the analysis because the algorithm accepts only identical primary criteria matches. Overall, our data loss was acceptable because of the high similarity that already existed at baseline. We were able to retain a large number of eyes (about 45%) as identical pairs of preoperative IOP and medication to focus on the preeminent questions of success in IOP and medication reduction. We found that FTO was as successful as TE with a similar reduction of IOP and medications. Both had a similar intervention and complication rate, notwithstanding numerical hypotony within the first six weeks after surgery. We observed a remarkably low rate of hyphema compared to ab interno trabeculectomy (Kaplowitz et al. 2016) that occurs when the IOP is at or below episcleral venous pressure allowing blood to reflux into the anterior chamber. This could indicate reduced patency of collector channels in advanced glaucoma that qualifies for filtering surgery, which has been observed ex vivo (Hann et al. 2014).

Trabeculotomy ab externo has been applied to adult POAG before but, compared to TE, was noted to have a lower success rate of 70% at one year, presumably due to a reapproximation or regeneration of the disrupted trabecular meshwork (Luntz & Livingston 1977). A study by Chihara et al. was in agreement with this, finding that a modified trabeculotomy ab externo lowers IOP to an average near 16 mmHg in a safe fashion (Chihara et al. 1993), by not as much as trabeculectomies. Ogawa et al. compared a nonpenetrating trabeculectomy with or without trabeculotomy (Ogawa et al. 2001) using a technique that was very similar to the one applied in our study except without MMC. Despite the absence of this antifibrotic, the authors achieved a 2-year IOP of 13 mmHg, not unlike our patients.

Our 2-year TE results match Kirwan et al.’s multicenter study of TE with an iridectomy in 428 eyes well (Kirwan et al. 2013). These eyes achieved an IOP of 12.4±4 mmHg and 80% did not have to use glaucoma medications anymore, slightly more than in our study population. The authors performed needling in 17% of patients, not unlike our rates and numerical hypotony during the first six months occurred in 7.2% of patients, which is in a similar range as our 9% in FTO and 2% in TE. Leakage was observed in 14% compared to 9% in our FTO and 2% in our TE. However, Kirwan et al.’s cataract surgery rate was 31%, much higher than our 1% in both FTO and TE, which might represent a difference in practice pattern or simply easier access to this elective procedure which is done as an outpatient surgery in the UK.

It is interesting to note that TEs in the TVT study were performed without an iridectomy (Gedde et al. 2005). In that study, IOP reduced from 25.6±5.3 mm Hg to 12.7±5.8 mm Hg at one year while the number of glaucoma medications declined from 3.0±1.2 to 0.5±0.9 (Gedde et al. 2007), which is also relatively similar to our results. An early complication rate of 37% was observed by these authors, primarily consisting of a shallow or flat anterior chamber in 20% and choroidal effusions in 10% (Gedde et al. 2012). In another study of TE without an iridectomy by Jea et al., IOP reduced from 26.3±10.9 mmHg to 10.2±4.1 mmHg at two years (Jea et al. 2012). The number of glaucoma medications decreased from 2.2±1.6 to 0.5±1.0. A complete success occurred in 76.6% at one year and 66.2% at two years, matching ours.

In all of these studies, trabeculectomy was performed as an outpatient patient procedure, a practice that emerged in the late 1980s (Beatty et al. 1996, Dobromyslov & Panina 1987) and became a standard for most types of eye surgeries and countries (Castells et al. 2001, Holland et al. 1992, Lleó et al. 2000, Wilson & Barr 1990). Even before the now-common use of antifibrotics, there was no significant difference in success or complication rates between inpatients and outpatients (Beatty et al. 1996), an observation that has been confirmed with antifibrotics as well (Lleó et al. 2000). In the country of our study, TE and FTO are only reimbursable as inpatient procedures. It has been argued that meticulous micromanagement after TE for several days may be associated with better long-term outcomes of TE. However, this hypothesis is challenged by the present study and by the findings of others (Beatty et al. 1996, Cortiñas et al. 2006, Dobromyslov & Panina 1987, Lleó et al. 2000). One could argue that the considerably lower cataract surgery rate in our data compared to Kirwan et al. (Kirwan et al. 2013) might indicate a higher threshold for cataract surgery. These would have typically also been done as inpatient procedures to better handle post-cataract surgery bleb care.

Our study was limited by its retrospective nature and nonrandomized design. The indication for postoperative interventions and length of hospitalization was at the discretion of the treating physicians and was not standardized for both groups. Despite reducing confounding through the exact matching of IOP and medications, other confounding factors might have contributed to small differences in early postoperative patient management, as reflected by the fact that FTO patients received more 5-FU injections. Although these patients had a higher rate of numerical hypotony they were hospitalized for a slightly shorter time and experienced results that were not significantly different.

In conclusion, our results are largely in line with other FTO and TE outpatient studies. Combining elements from both yields reasonable two-year rates of surgical success, postoperative complications, and interventions while avoiding an iridectomy.

## Data Availability

All relevant data is contained in the manuscript

## Supplemental Material

### Supplemental Figures

**Supplemental Figure 1:**
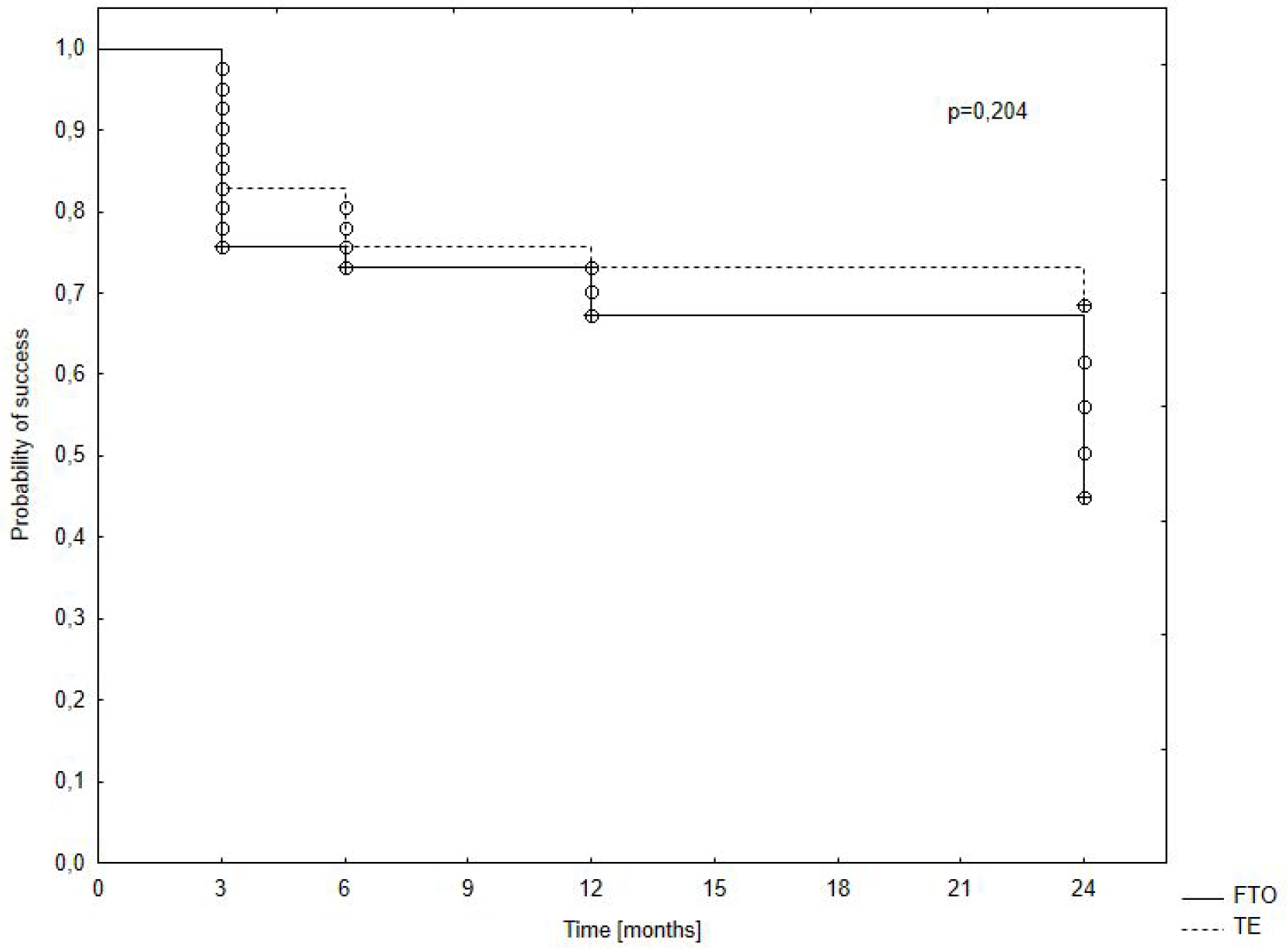
Kaplan-Meier curve for qualified success in FTO and TE.

**Supplemental Figure 2:**
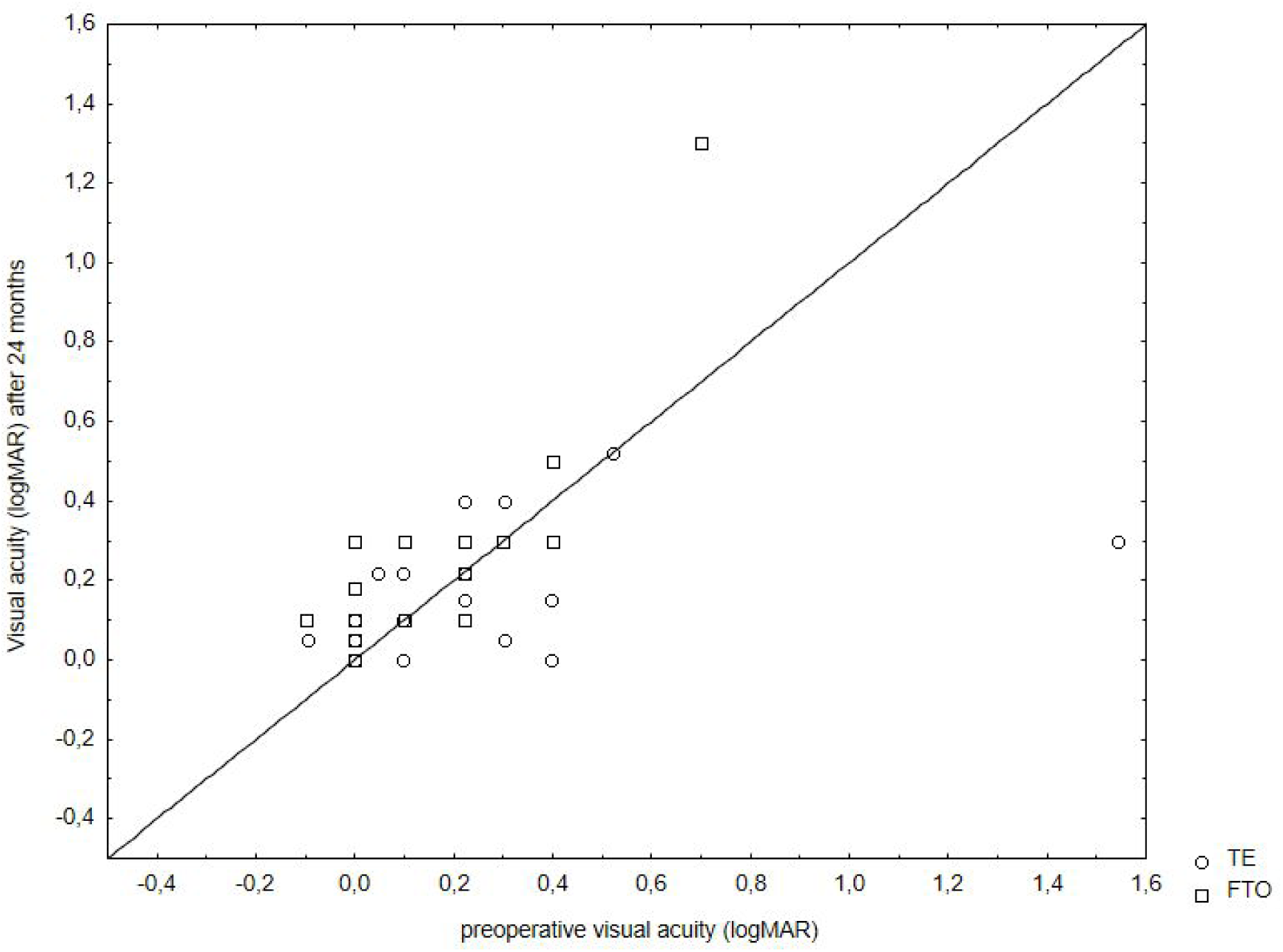
Scatter plots of visual acuity of both groups.

**Supplemental Figure 3:**
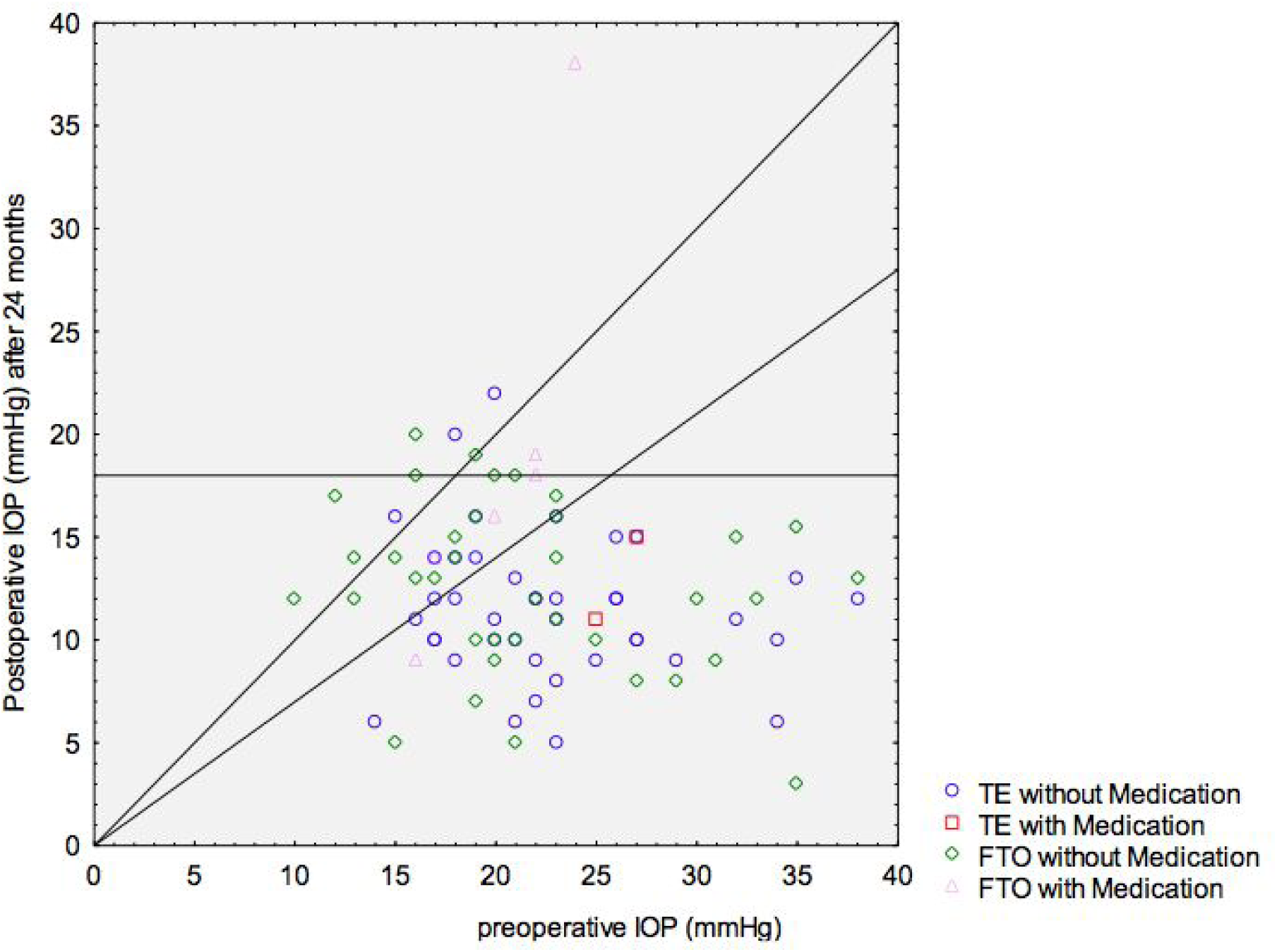
Scatter plots of IOP of both groups. Few patients required medications postoperatively.

## Notes

### Competing Interest Statement

The authors have declared no competing interest.

### Funding Statement

no external funding was received

## References

Abbas A, Agrawal P & King AJ (2018): Exploring literature-based definitions of hypotony following glaucoma filtration surgery and the impact on clinical outcomes. Acta Ophthalmol 96 : e285–e289.

Al Yousef Y, Strzalkowska A, Hillenkamp J, Rosentreter A & Loewen NA (2019, October 14): Comparison of a second-generation trabecular bypass (iStent inject) to ab interno trabeculectomy (Trabectome) by exact matching.

Aykan U, Bilge AH, Akin T, Certel I & Bayer A (2007): Laser suture lysis or releasable sutures after trabeculectomy. J Glaucoma 16 : 240–245.

Beatty S, Kheterpal S, Eagling EM & O’Neill EC (1996): Day-case trabeculectomies: Safety and efficacy. Acta Ophthalmol Scand 74 : 132–134.

Bindlish R, Condon GP, Schlosser JD & D’Antonio J (2002): Efficacy and safety of mitomycin-C in primary trabeculectomy: five-year follow-up. Ophthalmology.

Blackwell M, Iacus S, King G & Porro G (2009): Cem: Coarsened Exact Matching in Stata. Stata J 9 : 524–546.

Cairns JE (1968): Trabeculectomy. Preliminary report of a new method. Am J Ophthalmol 66 : 673–679.

Castells X, Alonso J, Castilla M, Ribó C, Cots F & Antó JM (2001): Outcomes and costs of outpatient and inpatient cataract surgery. Journal of Clinical Epidemiology. 23–29.

Chen CW, Huang HT, Bair JS & Lee CC (1990): Trabeculectomy with simultaneous topical application of mitomycin-C in refractory glaucoma. J Ocul Pharmacol 6 : 175–182.

Chihara E, Nishida A, Kodo M, Yoshimura N, Matsumura M, Yamamoto M & Tsukada T (1993): Trabeculotomy ab externo: an alternative treatment in adult patients with primary open-angle glaucoma. Ophthalmic Surg 24 : 735–739.

Cortiñas M, Martínez LL, Granados JM, Puerto N, Méndez M, Lizán-García M & López-Moya J (2006): [Results of an outpatient major surgery program in ophthalmology]. Arch Soc Esp Oftalmol 81 : 701–708.

de Barros DSM, Da Silva RS, Siam GA, et al. (2009): Should an iridectomy be routinely performed as a part of trabeculectomy? Two surgeons’ clinical experience. Eye 23 : 362–367.

Dobromyslov AN & Panina NB (1987): [Ambulatory care of patients with (surgically treated) glaucoma]. Oftalmol Zh 505–506.

Dvorak-Theobald G & Kirk HQ (1956): Aqueous pathways in some cases of glaucoma. Trans Am Ophthalmol Soc 53 : 301–15; discussion, 315–9.

Esfandiari H, Shazly TA, Waxman SA, Kola S, Kaplowitz KB, Brown EN & Loewen NA (2018): Similar Performance of Trabectome and Ahmed Glaucoma Devices in a Propensity Score-matched Comparison. J Glaucoma 27 : 490–495.

Esfandiari H, Taranum Basith SS, Kurup SP, et al. (2019): Long-term surgical outcomes of ab externo trabeculotomy in the management of primary congenital glaucoma. J AAPOS.

Esfandiari H, Taubenslag K, Shah P, et al. (2018, December 14): Two-year data comparison of ab interno trabeculectomy and trabecular bypass stenting using exact matching. J Cataract Refract Surg. 608–614.

Eslami Y, Mohammadi M, Khodaparast M, Rahmanikhah E, Zarei R, Moghimi S & Fakhraie G (2012): Sutureless tunnel trabeculectomy without peripheral iridectomy: a new modification of the conventional trabeculectomy. Int Ophthalmol 32 : 449–454.

Gedde SJ, Herndon LW, Brandt JD, Budenz DL, Feuer WJ & Schiffman JC (2012): Postoperative Complications in the Tube Versus Trabeculectomy (TVT) Study During Five Years of Follow-up. Am J Ophthalmol 153 : 804–814.e1.

Gedde SJ, Schiffman JC, Feuer WJ, Herndon LW, Brandt JD & Budenz DL (2007): Treatment Outcomes in the Tube Versus Trabeculectomy Study After One Year of Follow-up. American Journal of Ophthalmology. 9–22.e2.

Gedde SJ, Schiffman JC, Feuer WJ, Parrish RK 2nd, Heuer DK, Brandt JD & Tube Versus Trabeculectomy Study Group (2005): The tube versus trabeculectomy study: design and baseline characteristics of study patients. Am J Ophthalmol 140 : 275–287.

Grant WM (1958): Further studies on facility of flow through the trabecular meshwork. AMA Arch Ophthalmol 60 : 523–533.

Grehn F (1995): The value of trabeculotomy in glaucoma surgery. Curr Opin Ophthalmol 6 : 52–60.

Grehn F & Müller E (1990): Long-term results following preventive iridectomy. A retrospective study. Fortschr Ophthalmol 87 : 260–263.

Hann CR, Vercnocke AJ, Bentley MD, Jorgensen SM & Fautsch MP (2014): Anatomical Changes in Schlemm’s Canal and Collector Channels in Normal and Primary Open Angle Glaucoma Eyes Using Low and High Perfusion Pressures. Invest Ophthalmol Vis Sci.

Haubold B & Börsch-Haubold A (2017): Exact Matching. In: Haubold B & Börsch-Haubold A (eds.) Bioinformatics for Evolutionary Biologists: A Problems Approach. Cham: Springer International Publishing 47–67.

Holland GN, Earl DT, Wheeler NC, Straatsma BR, Pettit TH, Hepler RS, Christensen RE & Oye RK (1992): Results of inpatient and outpatient cataract surgery. A historical cohort comparison. Ophthalmology 99 : 845–852.

Honaker J, King G, Blackwell M & Others (2011): Amelia II: A program for missing data. J Stat Softw 45 : 1–47.

Hondur A, Onol M & Hasanreisoglu B (2008): Nonpenetrating glaucoma surgery: meta-analysis of recent results. J Glaucoma 17 : 139–146.

Jacobi PC & Krieglstein GK (1995): Trabecular aspiration. A new mode to treat pseudoexfoliation glaucoma. Invest Ophthalmol Vis Sci 36 : 2270–2276.

Jea SY, Francis BA, Vakili G, Filippopoulos T & Rhee DJ (2012): Ab interno trabeculectomy versus trabeculectomy for open-angle glaucoma. Ophthalmology 119 : 36–42.

Kaplowitz K, Bussel II, Honkanen R, Schuman JS & Loewen NA (2016): Review and meta-analysis of ab-interno trabeculectomy outcomes. Br J Ophthalmol 100 : 594–600.

Kayikcioglu OR, Emre S & Kaya Z (2010): Trabeculectomy combined with deep sclerectomy and scleral flap suture tension adjustment under an anterior chamber maintainer: a new modification of …. Int Ophthalmol.

King G (n.d.). Exact Matching.

Kirwan JF, Lockwood AJ, Shah P, et al. (2013): Trabeculectomy in the 21st century: a multicenter analysis. Ophthalmology 120 : 2532–2539.

Kostanyan T, Shazly T, Kaplowitz KB, Wang SZ, Kola S, Brown EN & Loewen NA (2017): Longer-term Baerveldt to Trabectome glaucoma surgery comparison using propensity score matching. Graefes Arch Clin Exp Ophthalmol 255 : 2423–2428.

Krasnov MM (1964): [SINUSOTOMY IN GLAUCOMA]. Vestn Oftalmol 77 : 37–41.

Krasnov MM (1968): Externalization of Schlemm’s canal (sinusotomy) in glaucoma. Br J Ophthalmol 52 : 157–161.

Lleó AVP, Moratal BJ, Pinós JPR, Hernández FJM, Navarro CP & Pallás CV (2000): Safety and efficacy of outpatient glaucoma surgery. Arch Soc Esp Oftalmol 75 : 535–540.

Luntz MH & Livingston DG (1977): Trabeculotomy ab externo and trabeculectomy in congenital and adult-onset glaucoma. Am J Ophthalmol 83 : 174–179.

Matlach J, Hipp M, Wagner M, Heuschmann PU, Klink T & Grehn F (2015): A comparative study of a modified filtering trabeculotomy and conventional trabeculectomy. Clin Ophthalmol 9 : 483–492.

McDonnell F, Dismuke WM, Overby DR & Stamer WD (2018): Pharmacological regulation of outflow resistance distal to Schlemm’s canal. Am J Physiol Cell Physiol 315 : C44–C51.

Neiweem AE, Bussel II, Schuman JS, Brown EN & Loewen NA (2016): Glaucoma Surgery Calculator: Limited Additive Effect of Phacoemulsification on Intraocular Pressure in Ab Interno Trabeculectomy. PLoS One 11 : e0153585.

Ogawa T, Dake Y, Saitoh AK, et al. (2001): Improved nonpenetrating trabeculectomy with trabeculotomy. J Glaucoma 10 : 429–435.

Ophir A (2001): Mini-trabeculectomy as initial surgery for medically uncontrolled glaucoma. Am J Ophthalmol 132 : 229–234.

Parikh HA, Bussel II, Schuman JS, Brown EN & Loewen NA (2016): Coarsened Exact Matching of Phaco-Trabectome to Trabectome in Phakic Patients: Lack of Additional Pressure Reduction from Phacoemulsification. PLoS One 11 : e0149384.

Pfeiffer N & Grehn F (1992): Improved suture for fornix-based conjunctival flap in filtering surgery. Int Ophthalmol 16 : 391–396.

Saul Sugar H (1961): Experimental Trabeculectomy in Glaucoma *. Am J Ophthalmol 51 : 623–627.

Schmier JK, Covert DW, Lau EC & Robin AL (2009): Trends in annual medicare expenditures for glaucoma surgical procedures from 1997 to 2006. Arch Ophthalmol 127 : 900–905.

Schwartz K & Budenz D (2004): Current management of glaucoma. Curr Opin Ophthalmol 15 : 119–126.

Shaarawy T, Sherwood M & Grehn F (2009): Guidelines on Design and Reporting of Glaucoma Surgical Trials. Kugler Publications.

Solus JF, Jampel HD, Tracey PA, Gilbert DL, Loyd TL, Jefferys JL & Quigley HA (2012): Comparison of limbus-based and fornix-based trabeculectomy: success, bleb-related complications, and bleb morphology. Ophthalmology 119 : 703–711.

Vuori M-L & Viitanen T (2001): ‘Scleral tunnel incision’-trabeculectomy with one releasable suture. Acta Ophthalmologica Scandinavica. 301–304.

Waxman S, Wang C, Dang Y, et al. (2018): Structure–Function Changes of the Porcine Distal Outflow Tract in Response to Nitric Oxide. Invest Ophthalmol Vis Sci 59 : 4886–4895.

Wilson D & Barr CC (1990): Outpatient and abbreviated hospitalization for vitreoretinal surgery. Ophthalmic Surg Lasers Imaging Retina.

